# SARS-CoV-2 seroprevalence in the municipality of São Paulo, Brazil, ten weeks after the first reported case

**DOI:** 10.1101/2020.06.29.20142331

**Authors:** Beatriz H. Tess, Celso F. H. Granato, Maria Cecília Goi Porto Alves, Maria Carolina Pintao, Edgar Rizzatti, Marcia C. Nunes, Fernando C. Reinach

## Abstract

A population-based household survey was performed to estimate the prevalence of IgM and IgG to SARS-CoV-2 in residents of six districts in São Paulo City, Brazil. Serum samples collected from 299 randomly-selected adults and 218 cohabitants (N=517) were tested by chemiluminescence immunoassay ten weeks after the first reported case. Weighted overall seroprevalence was 4.7% (95% CI 3.0-6.6%). The low seroprevalence suggests that most of this population could still be infected. Serial serosurveys were initiated aiming to monitor the progress of the ongoing pandemic throughout the entire city. This may help inform public health authority decisions regarding prevention and control strategies.

## Introduction

The spread of the infection with the severe acute respiratory syndrome coronavirus 2 (SARS-CoV-2) has been continuing worldwide since December 2019. In Brazil, the first laboratory-confirmed case was reported on February 26, 2020. The patient was a 61-year-old male resident in São Paulo City, who had returned from a visit to Lombardy (Italy) a few days previously [1]. On March 13^th^, 2020, the Brazilian Ministry of Health declared that community transmission had been established as a scaling up of cases of COVID-19 had been observed in multiple sites in the country and on March 24, the State of Sao Paulo declared a statewide quarantine. Highly urbanized with a population of approximately 11.6 million people and a high level of socioeconomic inequality, São Paulo City has been the epicenter of the COVID-19 epidemics in Brazil since then, registering 560 cases per 100.000 inhabitants, compared to the national figure of 249 cases per 100.000 as of 31 May 2020 [2].

Due to an extremely low testing capacity, epidemic projections and policies addressing COVID-19 in Brazil have been designed without robust data to inform epidemic parameters. Seroprevalence surveys are important for measuring the proportion of the general population who have been infected and have produced SARS-CoV-2 antibodies [3], which in turn, may confer protection (at least in the short term) against the infection.

We report results from a population-based household serosurvey planned to determine the prevalence of antibodies to SARS-CoV-2 in adults in six districts in São Paulo City, Brazil. We also investigated possible associations between selected risk factors for SARS-CoV-2 infection and the presence of SARS-CoV-2 antibodies.

## Methods

The design of this cross-sectional population-based study was designed informed by the World Health Organization (WHO) protocol for population-level COVID-19 antibody testing [4].

### Study population and study ethics

Six administrative districts of the São Paulo municipality were selected for the study because they were the most affected with COVID-19 according to official numbers of confirmed cases and suspected or confirmed deaths per 100.000 habitants as of April 23, 2020 [5]. Overall, 298.240 inhabitants aged 18 years old or more live in these districts [6].

All adults aged 18 years or older who were capable of understanding the information provided by the research teams were eligible to participate. A sample size of 500 households, one person per household, was planned to allow the estimation of the prevalence of seropositivity greater than 4% to be obtained under coefficients of variation of less than 30%. To allow for non-contact and refusals, 1049 households were randomly selected and surveyed.

Between May 4 and May 12 2020, field teams approached all households and randomly selected one member from each household. After providing written and verbal information on the study, the team invited the selected resident to participate. After obtaining informed consent from the participant, the team applied a structured questionnaire to gather information on sociodemographic characteristics and occurrence of COVID-19-related symptoms in the previous two weeks. Blood was collected by venipuncture following the interview. Other household members (cohabitants) who were interested in participating in the study, followed the same procedures. The study was approved by the Fleury Institutional Ethical Committee (CAAE 31032620.0.0000.5474).

### Specimen collection and laboratory tests

Samples were transported from the collection sites to the Serology Laboratory of Grupo Fleury where they were separated by centrifugation, aliquoted, and stored frozen until analysis, shortly after collection. All sera were tested for the presence of SARS-CoV-2 specific antibodies using the commercial kits “MAGLUMI IgM 2019-nCoV” and “MAGLUMI IgG 2019-nCoV” (chemiluminescence immunoassay-CLIA) supplied by Bioscience Co (China National Medical Products Administration: approval numbers 20203400183 (IgG) and 20203400182 (IgM); and Brazilian ANVISA RE 860 and 861/2020).

CLIA for IgG and IgM detection was developed based on a double-antibody sandwich immunoassay. The recombinant antigens containing the nucleoprotein and a peptide from the spike protein of SARS-CoV-2 were conjugated with FITC and immobilized on anti-FITC antibody-conjugated magnetic particles. Alkaline phosphatase-conjugated anti-human IgG and IgM was used as the detection antibody. The tests were conducted on an automated magnetic chemiluminescence analyzer (Axceed 260, Bioscience) according to the manufacturer’s instructions. The antibody titer was tested once per serum sample.

Detecting IgM and IgG antibodies against SARS-CoV-2 depends on the number of days after the onset of symptoms. The diagnostic performance of CLIA has been evaluated in the literature [7-11]. Although the sensitivity and specificity improve during the progression of infection, sensitivity for IgM is 100% and specificity 94.1% twenty days after symptoms onset. The IgG sensitivity and specificity, twenty days after the onset of symptoms is 100% and 99.5% respectively [7].

To determine the reference values, samples from patients diagnosed with the COVID-19 RT-PCR test and samples from control groups such as blood donors, employees without flu symptoms vaccinated in 2020, a panel of samples that tested positive for other non-SARS-CoV-2 respiratory viruses, and samples from carriers of other infectious diseases were used. The results of analyzing of these samples led to the following reference values: IgG: reagent >1.1 UA/mL, indeterminate 0.9 to 1.1 UA/mL, non-reagent <0.9 UA/mL; and IgM: reagent >1.0 UA/mL, indeterminate 0.7 to 1.0 UA/mL, non-reagent <0.7 UA/mL. Individuals who were reactive to either IgM or IgG were considered positive.

### Data analysis

Prevalence estimates were obtained for two groups separately: randomly-selected residents and cohabitants. Data were analyzed for both groups and were weighted according to the sampling design, which in turn, were adjusted considering response rates and post-stratification by age and sex, according to the district population structure in 2020 [6]. Prevalence estimates with 95% confidence intervals (CI) were calculated for selected characteristics, and frequency distributions of self-reported symptoms were compared between seropositive and seronegative individuals. Differences in proportions were assessed by the design-based F test. A p-value < 0.05 was considered to be statistically significant. Indeterminate results were included as negative in the analyses. STATA version 14 (StataCorp, College Station, TX, USA) was used for all statistical analyses.

## Results

Out of 771 contacted households, 299 (38.5% participation rate) randomly-selected residents agreed to participate in the study, as did 218 cohabitants. The seroprevalence for SARS-CoV-2 was 4.8% (95%CI 2.6-7.0%) for selected residents and overall seroprevalence, including test results for the 218 voluntary cohabitants, was 4.7% (95%CI 3.0-6.6%).

Of the 27 SARS-CoV-2 seropositive individuals, 20 were IgG positive/IgM negative, six were IgG negative/IgM positive, and one was IgG positive/IgM positive.

Socio-demographic characteristics of all 517 tested people stratified by SARS-CoV-2 seroprevalence estimates are shown in Table 1. Briefly, 54.9% were female, 52% aged between 18 and 44 years, 65% self-reported as being white and 46.4% had a university degree (16 years or more of schooling). Comparisons of seroprevalence show that people who reported brown and black skin color presented significantly higher SARS-CoV-2 seroprevalence compared to those with white skin. Nine or more years of schooling was associated with lower seroprevalence than low education level. Participants who reported one or more SARS-CoV-2-related symptoms in the past two weeks showed higher seropositivity than asymptomatic people, but this difference was not statistically significant (6.7% vs. 3.5%; P = .0801).

**Table 1.** SARS-CoV-2 seroprevalence, and 95% confidence interval, by selected population characteristics

| Characteristic | N of adults | Seroprevalence*<br>% | 95%CI | *p value* ** |
| --- | --- | --- | --- | --- |
| Overall | 517 | 4.7 | 3.0 - 6.6 | - |
| Gender |  |  |  |  |
| Female | 284 | 4.2 | 2.2 – 7.8 | 0.5821 |
| Male | 233 | 5.4 | 3.1 – 9.2 |  |
| Age (years) |  |  |  |  |
| 18-44 | 269 | 4.3 | 2.5 – 7.3 | 0.5916 |
| ≥ 45 | 248 | 5.2 | 3.1 – 8.5 |  |
| Skin colour/race*** |  |  |  |  |
| White | 335 | 3.3 | 1.9 – 5.6 | 0.0495**** |
| Brown and black | 147 | 8.6 | 4.7 – 15.0 |  |
| Asian and indigenous | 30 | 3.5 | 0.5 - 22.5 |  |
| Education (years of schooling) |  |  |  |  |
| ≤8 | 59 | 11.6 | 5.4 – 23.0 | 0.0251**** |
| 9-15 | 218 | 4.1 | 2.2 – 7.2 |  |
| ≥16 | 240 | 3.6 | 1.9 – 6.8 |  |
| SARS-CoV-2 related symptoms in the past 2 weeks |  |  |  |  |
| No symptom reported | 312 | 3.5 | 2.1 – 5.8 | 0.0801 |
| One or more symptoms reported | 205 | 6.7 | 3.8 – 11.7 |  |

\*corrected for sample design
\*\*design-based F test
\*\*\* missing data (n=5)
\*\*\*\* significant at 5%

Frequencies of self-reported symptoms during the two-week-period before the survey stratified by SARS-CoV-2 seroreactivity are shown in Table 2. Twelve (45.3%) of the 27 seropositive individuals did not report symptoms. SARS-CoV-2-seropositive individuals were significantly more likely to experience symptoms such as fatigue, diarrhea, ageusia, and anosmia than seronegative residents. None of the seropositive individuals reported previous diagnoses of infection.

**Table 2:** Frequency distribution, and 95% confidence interval, of self-reported symptoms in the last 14 days from date of survey by result of SARS-CoV-2 seroreactivity

| Symptoms* | Seronegative |  |  | Seropositive |  |  | p value** |
| --- | --- | --- | --- | --- | --- | --- | --- |
|  | n | % | 95%CI | n | % | 95%CI |  |
| Overall | 190 | 37.5 | 32.8 – 42.4 | 15 | 54.7 | 35.5 – 72.6 | 0.0801 |
| Fever | 10 | 2.1 | 0.9 - 4.8 | 2 | 5.6 | 1.2 - 22.0 | 0.2525 |
| Cough | 79 | 15.4 | 12.1 - 19.4 | 6 | 20.6 | 9.4 – 39.4 | 0.4556 |
| Shortness of breath | 24 | 4.7 | 2.7 – 7.9 | 2 | 5.6 | 1.2 – 22.0 | 0.7492 |
| Sore throat | 46 | 9.3 | 7.3 – 11.7 | 3 | 8.1 | 2.5 – 23.5 | 0.8226 |
| Rhinorrhea | 107 | 21.0 | 17.1 – 25.5 | 7 | 25.5 | 12.5 – 45.2 | 0.5827 |
| Fatigue | 32 | 5.8 | 3.6 – 9.3 | 5 | 20.3 | 8.5 – 41.2 | 0.0040*** |
| Myalgia | 46 | 8.9 | 6.2 – 12.7 | 6 | 18.8 | 8.3 – 37.1 | 0.0528 |
| Diarrhea | 28 | 5.5 | 3.7 – 8.0 | 4 | 13.2 | 5.3 – 29.0 | 0.0408*** |
| Ageusia | 14 | 2.9 | 1.6 – 5.3 | 6 | 18.4 | 7.1 – 39.7 | 0.0003*** |
| Anosmia | 15 | 3.0 | 1.7 – 5.4 | 6 | 18.4 | 7.1 – 39.7 | 0.0002*** |

\* single or multiple symptoms
\*\*design-based F test
\*\*\* significant at 5%

## Discussion

There are limited data available on the seroprevalence of SARS-CoV-2 infection in Brazil. In this study, an overall seroprevalence for anti-SARS-CoV-2 of 4.7% was observed. This estimate probably reflects the upper limit in the city of São Paulo since the selected districts were those with the highest number of cases and deaths in the city. One study, which used the Wondfo rapid antibody test, found a SARS-CoV-2 seroprevalence of 3.1% on May 17 [12]. Our findings were similar to this study, although seroprevalence estimates should be compared with caution, as the representativeness of the population covered, and laboratory testing methods differed between studies.

Besides assessing the magnitude of the pandemic in chosen areas of São Paulo City, we looked at possible associations between selected risk factors for SARS-CoV-2 infection and the presence of SARS-CoV-2 antibodies. Previous reports have linked SARS-CoV-2 infections with no or minor symptoms [1,13,14]. We found results in the same direction, 45.3% of seropositive individuals reported no symptoms in the two weeks before the survey. Similar to studies on the frequency of SARS-CoV-2-related symptoms, anosmia, ageusia, fatigue and diarrhea were symptoms associated with seropositivity [13,14]. It is noteworthy that SARS-CoV-2-related clinical symptoms and signs remain poorly documented in Brazil.

The strengths of the current research are that it is the first household population-based serosurvey in Brazil using a chemiluminescence assay, a very precise technique to detect immunoglobulins against SARS-CoV-2. This test together with the probabilistic sampling design provided a clear picture of the extent of human exposure to SARS-CoV-2 in the studied regions.

There are some limitations to the study. First, since it was a cross-sectional study, temporal associations cannot be established and data are lacking on variables related to the epidemiology of SARS-CoV-2, such as the history of exposure and severity of symptoms. Second, refusal bias may have been introduced as a result of non-response, which occurred mainly due to non-cooperation by the managers of some residential blocks and to residents’ unwillingness to donate blood. Third, we have not included children or young people under 18 years due to logistic restrictions. Last, the statistical power may have been insufficient to detect differences between subgroups. However, we believe that these issues have not affected our findings significantly, considering that our results are consistent with several worldwide reports that showed low prevalence of SARS-CoV-2 infection in the general population even in highly affected regions [12,13,15].

In conclusion, the overall exposure to SARS-CoV-2 in the general population in the research areas is low, indicating that a significant proportion of this population could still be infected. Following these initial results, a series of six monthly point-prevalence serosurveys, using similar methodology and the same testing method, has been initiated. By monitoring the SARS-CoV-2 seroprevalence in a population-representative sample, as well as the ongoing pandemic in São Paulo City, high-risk groups and areas may be identified which will help guide public health actions.

## Data Availability

The datasets analyzed during the current study are available from the corresponding author on reasonable request.

## Acknowledgments

We thank study participants whose cooperation made the study possible. We thank Pedro Passos, Guilherme Passos, Carlos Marinelli, A. Hernandez, A. Resende, D. Freitas, Fernando Pieroni, Jefferson Tolentino, Assis, L. S. Silva, V.A. Frascini, Willian Malfatti, Rosi Rosendo, Helio Neves, Sofia Reinach and Adriano Borges Costa for their help. We are grateful for the support of Dr Carolina dos Santos Lazari.

